# OpenSlideFM: A Computationally Efficient Multi-Scale Foundation Model for Computational Pathology

**DOI:** 10.64898/2025.12.09.25341769

**Authors:** Sanwal Ahmad Zafar, Wei Qin, Liu Chengliang, Areeba Ali Khan, Alina Nazir, Farhan Khalid, Muhammad Salman Faisal

**Affiliations:** School of Mechanical Engineering, Shanghai Jiao tong University, Shanghai, China; Services Institute of Medical Sciences, Pakistan; Tameside and Glossop Integrated Care NHS Foundation Trust, Manchester, UK; Union Hospital, Terre Haute, Indiana, USA; Stephenson Cancer Center, OU Health University, Oklahoma Medical Centre, Oklahoma, USA

**Keywords:** computational pathology, foundation models, whole-slide imaging, multi-scale learning, self-supervised learning, cancer classification

## Abstract

**Background:** Computational pathology increasingly relies on foundation models pre-trained on large-scale histopathology datasets, but existing models require substantial computational resources that limit accessibility for resource-constrained institutions. We present OpenSlideFM, a computationally efficient multi-scale foundation model that balances performance with practical deployment requirements.

**Methods:** We developed a dual-scale transformer architecture that simultaneously processes high-resolution (0.5 μm/pixel) and low-resolution (2.0 μm/pixel) features to capture both cellular morphology and tissue architecture. The model was pre-trained using self-supervised learning on 20,000 whole-slide images from The Cancer Genome Atlas spanning 31 cancer types and 10,795 patients (Table 1). We validated OpenSlideFM across four independent tasks: pan-cancer classification, lymph node metastasis detection, pathological staging, and prostate cancer grading.

**Results:** OpenSlideFM achieved 81.21% accuracy on 31-class pan-cancer classification, macro-AUROC of 98.65%. Multi-scale architecture significantly outperformed single-scale variants, with 2.35% absolute improvement over high-resolution alone. External validation demonstrated robust generalization: 77.4% AUROC for lymph node metastasis detection, 0.254 quadratic kappa for multi-center pathological staging, and 0.826 quadratic kappa for prostate cancer grading. The model requires only 35 million parameters and trains on a single consumer-grade workstation with NVIDIA GeForce RTX 4090 GPU (24 GB VRAM, 16-core CPU, 384 GB RAM), enabling accessible deployment compared to 300-1850 million parameters and datacenter GPU requirements for existing foundation models.

**Conclusions:** OpenSlideFM demonstrates that computationally efficient foundation models can achieve competitive performance across diverse histopathology tasks while maintaining practical deployment feasibility. The multi-scale architecture provides complementary information from cellular and tissue levels, and the reduced computational requirements democratize access to foundation model capabilities for resource-constrained medical institutions.

## 1. INTRODUCTION

Histopathological examination of tissue samples remains the gold standard for cancer diagnosis, but manual microscopic assessment faces challenges including inter-observer variability, labor intensity, and limited scalability. The advent of whole-slide imaging (WSI) technology has digitized pathology workflows, enabling the application of artificial intelligence to automate and augment diagnostic tasks. Recent advances in computational pathology have been driven by foundation models, large-scale neural networks pre-trained on diverse histopathology datasets using self-supervised learning.

Foundation models such as UNI [1], Virchow [2], and CONCH [3] have demonstrated remarkable performance across multiple computational pathology tasks. UNI is pretrained using **>100 million** image tiles from **>100,000 diagnostic WSIs** (**>77 TB**) across ∼20 tissue types [1]. Virchow utilized 1.5 million whole-slide images from 100,000 patients [2], while Virchow2-Giant scales to 1.85 billion parameters. However, these models require substantial computational resources, specifically high-end datacenter GPUs with extensive memory, that may be prohibitive for resource-constrained medical institutions, community hospitals, and low-resource settings. The hardware investment required can be challenging, as training and deploying models like UNI (307M parameters) or Virchow2 (1.85B parameters) requires NVIDIA Datacenter-class GPU or equivalent memory capacity. This hardware barrier limits the democratization of AI-driven pathology capabilities and creates disparities in access to advanced diagnostic tools.

Beyond computational constraints, most existing foundation models process patches at a single fixed resolution, potentially missing complementary information across scales. Pathologists naturally examine tissue at multiple magnifications, alternating between high-power fields for cellular detail and low-power fields for architectural patterns. High resolution captures nuclear morphology, cellular organization, and fine-grained patterns essential for subtype discrimination, while low resolution provides spatial context, tissue architecture, and structural relationships important for understanding tumor microenvironment and growth patterns.

This work presents OpenSlideFM, a computationally efficient foundation model that addresses the accessibility gap while maintaining competitive performance. Our specific contributions include: (1) a multi-scale architecture, a dual-resolution framework (0.5 μm + 2.0 μm) that simultaneously captures cellular morphology and tissue architecture (Figure 1A), achieving 2.35% absolute improvement over single-scale baselines (p < 0.001) (Supplementary Table 2A); (2) computational efficiency through a 35-million-parameter model deployable on consumer-grade GPUs (∼12 GB inference; ∼22 GB training; 24 GB recommended), representing 8.8× fewer parameters than UNI and 53× fewer than Virchow2-Giant (Table 3); (3) comprehensive validation across four diverse clinical tasks spanning different organs (breast, lung, prostate, kidney), objectives (classification, detection, grading), and datasets (32,014 total slides), demonstrating robust generalization (Table 1, Table 2); (4) systematic ablations rigorously quantifying the impact of architectural choices (scale, aggregation, token budget) with statistical significance testing (Supplementary Tables 2A-C); and (5) a commitment to open science through releasing model weights and code to facilitate reproducibility and broader adoption in the computational pathology community.

**Figure 1.**
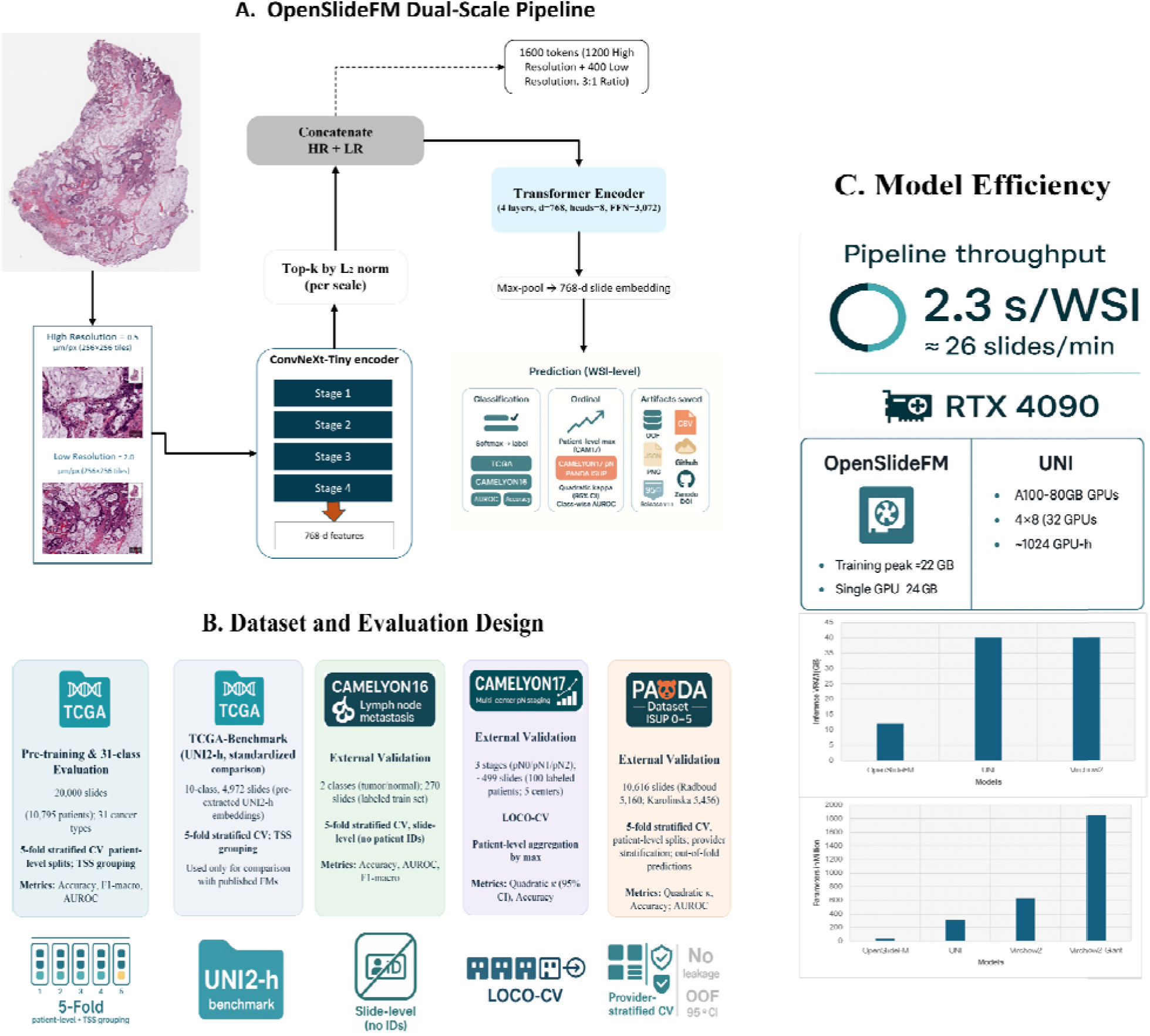
OpenSlideFM architecture, evaluation design, and computational efficiency. **(A)** Dual-scale feature extraction pipeline processes patches at 0.5 μm/pixel (high resolution) and 2.0 μm/pixel (low resolution), concatenates features, and applies transformer aggregation for slide-level prediction. **(B)** Comprehensive evaluation across pre-training (TCGA), benchmark comparison (UNI2-h), and external validation datasets (CAMELYON16/17, PANDA) using stratified cross-validation strategies. **(C)** Computational efficiency: 2.3 seconds per slide throughput, single consumer GPU deployment (RTX 4090, 24 GB), and 8-53× parameter reduction compared to existing foundation models.

**Table 1.** Multi-Dataset Evaluation Design.

| Dataset | Task | Classes/Stages | Slides/Patients | Validation Method | Primary Metrics |
| --- | --- | --- | --- | --- | --- |
| **TCGA-Full** | Pan-cancer classification | 31 cancer types | 20,000 slides (10,795 patients) | 5-fold stratified CV, Patient-level splits, TSS grouping | Accuracy, F1 Score, AUROC |
| **TCGA-Benchmark (UNI2-h)** | 10-class | 10 cancer types (ACC, BRCA-IDC, COAD, DLBC, GBM, HNSC, KIRC, LUAD, SKCM, UCEC) | 4,972 slides | 5-fold stratified CV, TSS grouping | Accuracy, F1 Score, AUROC |
| **TCGA (OpenSlideFM)** | 10-class | 10 cancer types (ACC, BRCA-IDC, COAD, DLBC, GBM, HNSC, KIRC, LUAD, SKCM, UCEC) | 4,972 slides | 5-fold stratified CV Patient-level splits, TSS grouping |  |
| **CAMELYON16** | Lymph node metastasis detection | 2 classes (tumor/normal) | 270 slides (labeled train set) | 5-fold stratified CV, slide-level splitting (no patient IDs) | Accuracy, AUROC, Precision, F1-score |
| **CAMELYON17** | Multi-center pN staging | 3 stages (pN0, pN1, pN2) | 499 slides (100 labeled patients across 5 centers) | Leave-one-center-out CV (LOCO-CV) on labeled set; patient-level aggregation by max | Quadratic $\kappa$ 95% CI, Accuracy |
| **PANDA** | Prostate cancer Gleason | 6 ISUP grades (0-5) | 10,616 slides (2 providers: Radboud 5,160, | 5-fold stratified CV, Patient-level splits, | Quadratic $\kappa$, Accuracy, |
|  | grading |  | Karolinska<br>5,456) | Provider<br>stratification,<br>Out-of-fold<br>predictions | AUROC |

**Table 2.** Primary Performance Results Across All Datasets.

| Dataset | Task Description | Primary Metric | Result (Mean ± SD) | 95% Confidence Interval | Secondary Metrics |
| --- | --- | --- | --- | --- | --- |
| **TCGA-Full** | 31-cancer-typeclassification | Accuracy | **81.21%** ± **0.56%** | [80.35%, 82.08%] | AUCOR: 98.65% ± 0.09% Bal-Acc: 76.88% ± 1.33% F1-macro: 75.54% ± 1.15% |
| **TCGA-Benchmark (OpenSlideFM)** | 10-class benchmark (4972 Slides) | Accuracy | **81.8%** ± **1.9%** | [79.1%, 84.2%] | AUCOR: 0.929 ± 0.027 |
| **TCGA-Benchmark (UNI1-h)** | 10-class foundation model comparison | Accuracy | **92.04%** ± **0.78%** | [90.77%, 93.21%] | AUCOR: 99.38% ± 0.11% Bal-Acc: 89.52% ± 0.92% F1-macro: 88.67% ± 1.05% |
| **CAMELYON16** | Lymph node metastasis detection | AUC | **0.774** ± **0.045** | [0.697, 0.810] | Accuracy: 68.8% ± 4.2% F1-score: 56.3% ± 5.8% Avg Precision: 68.1% ± 4.1% |
| **CAMELYON17** | Multi-center pN staging (LOCO-CV) | Quadratic $\kappa$ | **0.254** ± **0.092** | [0.062, 0.429] | Mean CV $\kappa$: 0.204 ± 0.091 Accuracy: |
| | | | | | 52.3% ± 8.7% Per-stage AUC: 0.60-0.72 |
| **PANDA** | Prostate ISUP grading (6 classes) | Quadratic $\kappa$ | **0.826 ± 0.008** | [0.810, 0.842] | Accuracy: 67.0% ± 0.9% Macro-AUROC: 0.910 ± 0.005 |

**Table 3.** Comparison with Foundation Models.

| Model | Parameters | Pre-Training Data | Pre-Training Algorithm | TCGA Performance† | CAMELY ON16 AUC | PANDA Quadratic κ | Deployment Accessibility |
| --- | --- | --- | --- | --- | --- | --- | --- |
| **OpenSlideFM** | **35M**(28M backbone + 7M aggregator) | 20K WSIs, 31 cancer types, 10,795 patients | Self-supervised (BYOL + MFR) | **81.21%**(31-class, 20K slides) | **0.774** | **0.826** | **High** Consumer-grade hardware |
| **UNI** | 307M | 100M images, 100,402 WSIs, 20 tissue types | Self-supervised (DINOv2) | **92.0%** (10-class, 4,972 slides) | 79.5% | 0.839(reported) | Medium Datacenter GPU |
| **Virchow2** | 632M | 3.1M WSIs | Self-supervised | ~90% (10-class, | 81.2% | 0.847(reported) | Medium Datacent |
|  |  |  | (DINOv2) | reported) |  |  | er GPU |
| **Virchow2-Giant** | 1.85B | 1.5M WSI 100K patients 3.1M slides total | Self-supervised(DINOv2) | ~91%(10-class, reported) | - | Not reported | Low High-end datacenter |
| **CONCH** | 307M | 1.17M image-text pairs Diverse sources | Vision-language contrastive | ~89%(10-class, reported) | 78.3% | 0.813(reported) | Medium Datacenter GPU |

## 2. MATERIALS AND METHODS

### DATASETS

We utilized one primary pre-training dataset and three external validation datasets to assess model performance across diverse histopathology tasks (Table 1, Figure 1B). The Cancer Genome Atlas (TCGA) provided 20,000 hematoxylin and eosin (H&E) stained whole-slide images from 10,795 patients spanning 31 cancer types [4]. Slides were obtained from multiple tissue source sites across the United States, ensuring geographic and institutional diversity. Cancer types included breast invasive carcinoma (BRCA, n=1,092 patients), kidney renal clear cell carcinoma (KIRC, n=526), lung adenocarcinoma (LUAD, n=513), head-neck squamous cell carcinoma (HNSC, n=513), brain lower grade glioma (LGG, n=510), thyroid carcinoma (THCA, n=503), lung squamous cell carcinoma (LUSC, n=497), prostate adenocarcinoma (PRAD, n=490), skin cutaneous melanoma (SKCM, n=464), ovarian serous cystadenocarcinoma (OV, n=422), and 21 additional cancer types. Whole-slide images were acquired at 40× magnification (0.25 μm/pixel) using Aperio scanners. We applied tissue segmentation to remove background regions and extracted non-overlapping patches at two resolutions: 256×256 pixels at 0.5 μm/pixel (high resolution, 128 μm tissue area) and 256×256 pixels at 2.0 μm/pixel (low resolution, 512 μm tissue area). Patches with tissue coverage below 50% were excluded. We performed patient-level stratified 5-fold cross-validation to prevent data leakage, allocating approximately 80% of patients to training and 20% to validation. The cross-validation was repeated across 3 independent runs with seeds 42, 123, 456 for a total of 15 evaluations (5 folds × 3 runs). Tissue source site (TSS) codes were used as grouping variables to account for site-specific batch effects. To enable fair comparison with published foundation models, we additionally evaluated pre-extracted UNI2-h features on a standard 10-cancer-type benchmark subset (4,972 slides) following the evaluation protocol established by Chen et al. [1]. This subset includes ACC (n=227), BRCA-IDC (n=838), COAD (n=442), DLBC (n=44), GBM (n=858), HNSC (n=472), KIRC (n=519), LUAD (n=531), SKCM (n=475), and UCEC (n=566) (Supplementary Table 1B). Pre-extracted embeddings were obtained from the Mahmood Lab public repository. UNI was pre-trained on independent data that explicitly excluded TCGA, CPTAC, PAIP, CAMELYON, PANDA, and other public benchmarks [1].

The CAMELYON16 challenge dataset [5] consists of 400 H&E-stained sentinel lymph node sections from two medical centers in the Netherlands, divided into a training set (270 slides: 111 tumor, 159 normal) and a test set (129 slides: 30 tumor, 100 normal) (Supplementary Table 1B). Slides with micrometastases or macrometastases were labeled as positive, while slides without metastatic involvement were labeled as negative. We used the 270 labeled training slides for 5-fold stratified cross-validation. The 129 official test slides were held out for unlabeled external checks only. Slides were scanned at 40× magnification (0.25 μm/pixel). The CAMELYON17 challenge [6] addresses multi-center generalization for pathological lymph node (pN) staging, including 200 patients from five medical centers in the Netherlands with approximately 5 H&E slides per patient (1000 total slides: 500 training, 500 test) (Supplementary Table 1C). Patients were classified into three pN stages based on 2009 AJCC/UICC TNM staging: pN0 (no lymph node metastasis), pN1 (micrometastasis or 1-3 positive nodes), and pN2 (≥4 positive lymph nodes). We performed leave-one-center-out cross-validation on the 100 labeled patients (499 slides): train on four centers and validate on the held-out center. Slides were scanned at 40× magnification.

The Prostate cANcer graDe Assessment (PANDA) challenge dataset [7] comprises 10,616 digitized H&E-stained prostate biopsies from two centers: Radboud University Medical Center (n=5,160, 48.6%) and Karolinska Institute (n=5,456, 51.4%) (Supplementary Table 1D). Slides were annotated with International Society of Urological Pathology (ISUP) grades ranging from 0 (benign) to 5 (most aggressive). The distribution was: Grade 0 (n=2,892, 27.2%), Grade 1 (n=2,666, 25.1%), Grade 2 (n=1,343, 12.6%), Grade 3 (n=1,242, 11.7%), Grade 4 (n=1,249, 11.8%), and Grade 5 (n=1,224, 11.5%). Radboud slides were scanned at 20× magnification (0.5 μm/pixel), while Karolinska slides were scanned at 40× magnification (0.25 μm/pixel). We performed 5-fold stratified cross-validation with patient-level splitting.

### MODEL ARCHITECTURE

We employed ConvNeXt-Tiny [8] as the patch-level feature extractor, providing a balance between representational capacity and computational efficiency. ConvNeXt-Tiny contains 28 million parameters and processes 256×256-pixel patches through four hierarchical stages with depthwise convolutions, producing 768-dimensional feature vectors. We extracted features at two resolutions: high resolution (0.5 μm/pixel) captures cellular morphology, nuclear features, and fine-grained patterns, yielding 2,000-5,000 patches per WSI; low resolution (2.0 μm/pixel) captures tissue architecture, spatial relationships, and structural organization, yielding 125-315 patches (16× reduction in patch count) **(Figure 2A)**.

**Figure 2.**
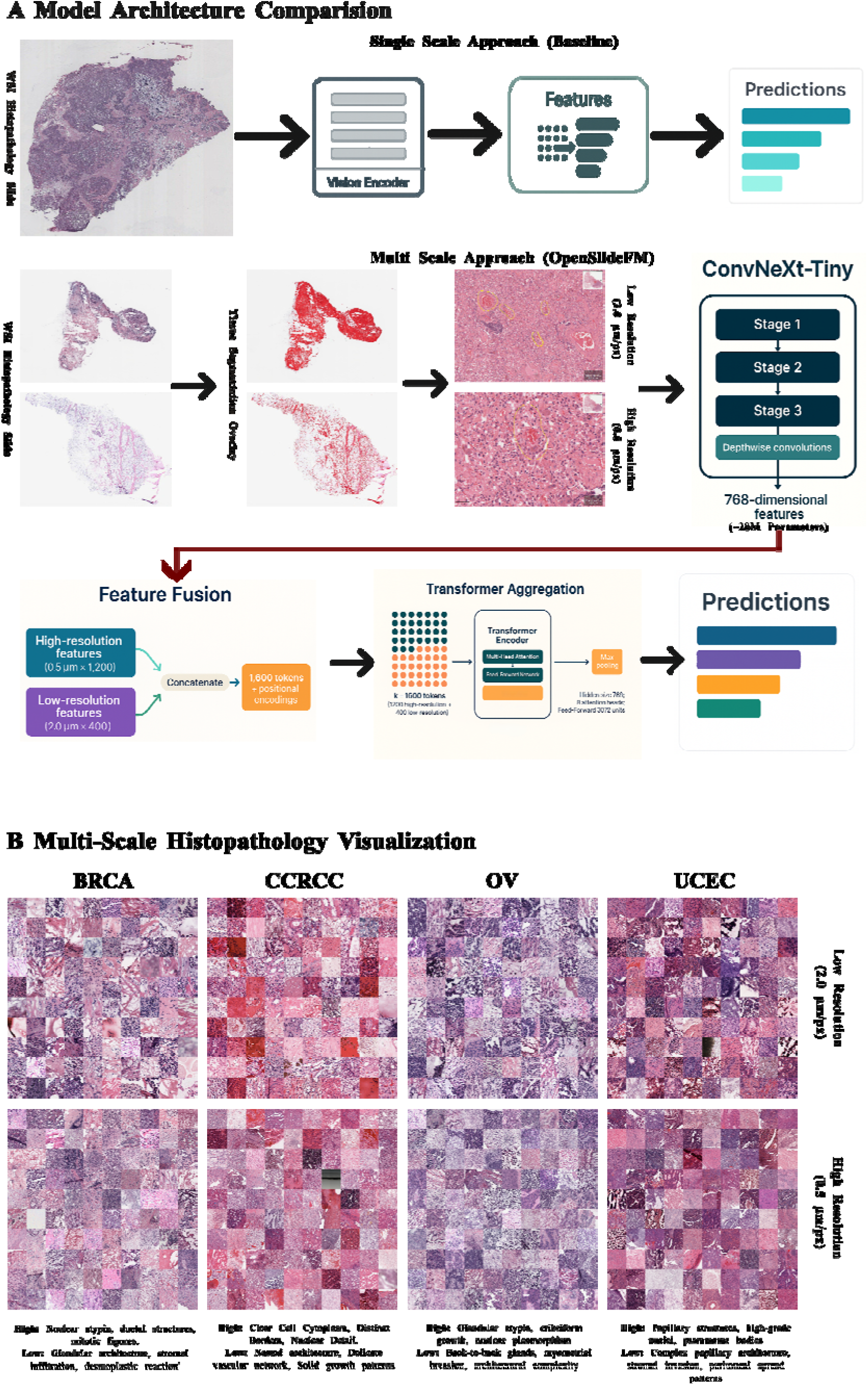
Multi-Scale Architecture and Histopathology Visualization. **(A)** Architecture comparison. Single-scale baseline (top) processes WSIs at fixed resolution. OpenSlideFM (bottom) extracts features at dual resolutions, 0.5 μm/pixel for cellular morphology and 2.0 μm/pixel for tissue architecture, using ConvNeXt-Tiny with transformer-based cross-scale fusion. **(B)** Multi-scale examples across cancer types. High magnification (top, 0.5 μm/pixel) captures nuclear features and cellular details; low magnification (bottom, 2.0 μm/pixel) captures architectural patterns and stromal organization. Representative TCGA samples: BRCA (breast invasive carcinoma), CCRCC (clear cell renal cell carcinoma), OV (ovarian serous cystadenocarcinoma), UCEC (uterine corpus endometrial carcinoma).

To maintain computational tractability while preserving diagnostic information, we implemented adaptive token selection. Given a whole-slide image, we extracted all patches at both resolutions, then selected the top-k patches by L2 norm of their feature vectors. The token budget was allocated as 75% high-resolution (1,200 tokens) and 25% low-resolution (400 tokens), totaling 1,600 tokens, prioritizing cellular detail while ensuring adequate representation of tissue architecture (Supplementary Table 2C). Selected tokens from both resolutions were concatenated and processed through a 4-layer transformer encoder (hidden dimension: 768, attention heads: 8, feedforward dimension: 3,072) that learns cross-scale interactions and generates slide-level representations [9] **(Figure 2A)**. We applied max pooling over the transformer output to produce a final 768-dimensional slide embedding, selected based on ablation studies showing comparable performance to mean pooling with better computational properties (Supplementary Table 2B).

For classification tasks (TCGA, CAMELYON16), we used logistic regression with L2 regularization (C=1.0) on slide embeddings. For ordinal grading tasks (CAMELYON17 pN staging, PANDA ISUP grading), we employed ordinal regression with cumulative link models. For PANDA, we additionally implemented multiple instance learning (MIL) with attention-based aggregation [10] to handle the extreme variability in biopsy size (100-50,000 patches per slide).

### SELF-SUPERVISED PRE-TRAINING

We pre-trained the feature extraction backbone on the full TCGA dataset (20,000 slides, 31 cancer types) using two complementary self-supervised objectives: Bootstrap Your Own Latent (BYOL) [11], a momentum-based contrastive learning framework that learns representations by predicting augmented views of patches using EMA coefficient 0.996, predictor hidden dimension 2,048, and training for 4 epochs with batch size 3 slides and gradient accumulation over 2 steps (effective batch size 6 slides); and Masked Feature Reconstruction (MFR), a generative objective that masks 50% of patch tokens and reconstructs their features using a decoder network to encourage learning spatially coherent representations and understanding of tissue context. During pre-training, tissue tiles were processed through the ConvNeXt-Tiny encoder in batches of 128 tiles for computational efficiency, distinct from the slide-level batch size used during overall model training.

We applied stochastic augmentations to each patch during pre-training: random horizontal and vertical flipping (p=0.5), random rotation (±90°, p=0.25), color jittering (brightness ±0.2, contrast ±0.2, saturation ±0.1, hue ±0.05), Gaussian blur (σ=0.1-2.0, p=0.5), and stain normalization using Macenko method [12]. Training was conducted on a single workstation with NVIDIA GeForce RTX 4090 (24 GB VRAM), 384 GB RAM, and 16-core CPU, using AdamW optimizer [13] (β =0.9, β =0.999, weight decay=1e-4), learning rate 1.5×10 with cosine annealing and 500-step warmup, batch size of 3 slides with gradient accumulation over 2 steps (effective batch size 6 slides), token budget of 1,200 @ 0.5μm + 400 @ 2.0μm = 1,600 tokens/slide yielding 9,600 tokens per optimization step, training duration of 4 epochs (∼72 hours), and mixed precision FP16 with gradient scaling for memory efficiency (Supplementary Table 3).

### ARCHITECTURE DESIGN ABLATIONS

We conducted comprehensive ablation studies to optimize OpenSlideFM’s architecture and validate design decisions using the full TCGA dataset (20,000 slides, 31 cancer types) with 5-fold stratified cross-validation, patient-level splitting, and tissue source site grouping (Supplementary Tables 2A-C).

To quantify the contribution of multi-scale feature extraction, we tested three variants: 0.5μm only (high-resolution patches only, 1,600 tokens), 2.0μm only (low-resolution patches only, 1,600 tokens), and both scales (dual-resolution, 1,200 high-res + 400 low-res tokens) (Supplementary Table 2A). The multi-scale architecture achieved 82.32% ± 0.29% accuracy, significantly outperforming single-scale variants: 79.97% ± 0.20% for high-resolution only (Δ=-2.35%, p<0.001) and 77.28% ± 0.56% for low-resolution only (Δ=-5.04%, p<0.001), demonstrating complementary information from cellular morphology and tissue architecture (Figure 2B). All ablations used 5-fold cross-validation repeated across 3 seeds (42, 123, 456); significance was computed with a paired t-test on the 15 fold-wise scores per variant.

We compared simple pooling methods against heuristic hard selection mechanisms by testing mean pooling (average of patch features, O(1) complexity), max pooling (maximum values across patch features, O(1) complexity), and top-k pooling (select top 15% patches by feature norm, O(n log k) complexity) (Supplementary Table 2B). Concatenated patch embeddings from both scales without transformer aggregation were directly pooled into slide-level representation, followed by logistic regression classifier. Mean pooling achieved 62.48% ± 0.91% accuracy, max pooling achieved 63.99% ± 0.74% (Δ=+1.51%, p=0.089), and top-k pooling achieved 56.17% ± 1.00% (Δ=-6.31%, p<0.001). Simple pooling methods performed comparably, while heuristic hard selection significantly underperformed. The full model with transformer aggregation achieves 81.21% accuracy, an 18.7% absolute improvement over pooling-only methods, demonstrating the value of learned cross-scale interactions (Supplementary Table 2B). †Note: The ablation study reports 82.32% ± 0.29% using a single random seed (42) for controlled comparison. Final results report 81.21% averaged across 3 independent runs with different seeds (42, 123, 456), providing 15 total cross-validation evaluations (5 folds × 3 runs) for robust performance estimation.

To determine optimal token budget balancing performance and computational cost, we tested 100, 250, 500, 750, 1000, 1600, 2000 tokens with fixed 3:1 ratio (high-resolution:low-resolution) (Supplementary Table 2C). Performance improved monotonically from 62.7% (100 tokens) to 82.3% (1,600 tokens), with marginal decline at 2,000 tokens (81.9%). The 1,600-token configuration represents optimal balance, while 1,000 tokens provide efficient alternative at 67.8% accuracy with 37.5% computational savings. We recommend 1,600 tokens for high-performance applications and 750-1,000 tokens for resource-constrained settings achieving 66-68% accuracy.

### EVALUATION PROTOCOL

For TCGA, we performed 5-fold stratified cross-validation with patient-level splitting using tissue source site (TSS) codes as grouping variable in stratified group k-fold to prevent leakage from slides of the same patient or institution. Each fold allocated 80% training and 20% testing, repeated across 3 independent runs with different random seeds (42, 123, 456). For CAMELYON16, we used the **270-slide CAMELYON16 training set** for 5-fold stratified cross-validation. The **130-slide official test set** was held out and **not** used for training/model selection. For CAMELYON17, we used leave-one-center-out cross-validation (LOCO-CV) across 5 medical centers, training on four centers (∼80 patients) and validating on one held-out center (∼20 patients) per fold on the 100 labeled training patients (≈499 slides), with patient-level predictions aggregated from slide-level probabilities using max operator. For PANDA, we performed 5-fold stratified cross-validation with patient-level splitting and provider stratification (Radboud vs Karolinska) to ensure balanced representation, using out-of-fold predictions for final evaluation.

For classification tasks (TCGA, CAMELYON16), primary metrics were accuracy and area under receiver operating characteristic curve (AUROC) [14], with secondary metrics including balanced accuracy, macro-averaged F1-score [15], and macro-averaged precision and recall. For ordinal tasks (CAMELYON17, PANDA), primary metric was quadratic weighted Cohen’s kappa (κ) [16] accounting for ordinal relationships, with secondary metrics including accuracy and class-wise AUROC (one-vs-rest). Statistical significance was assessed using 95% confidence intervals computed from 1,000 bootstrap replicates [17] with replacement and paired t-tests for comparing methods across CV folds with Bonferroni correction for multiple comparisons.

Training was conducted on a single workstation with NVIDIA GeForce RTX 4090 (24 GB VRAM), 384 GB RAM, and 16-core CPU (AMD64, 3.6 GHz), requiring 72 hours for pre-training (4 epochs) with peak ∼22 GB GPU memory usage (Supplementary Table 3). Inference on the same hardware achieved 2.3 seconds per whole-slide image with single-stream processing, using ∼12 GB for single-stream and ∼16-22 GB with moderate concurrency (24 GB card recommended for 8 slides simultaneously). Single-stream latency is 2.3 seconds per WSI (∼26 slides/min), while 2-4 concurrent decode/forward streams on one RTX 4090 achieve ∼37 slides/min sustained throughput.

Implementation used PyTorch 2.5.1 [18] with CUDA 12.1, Python 3.11.1 on Windows 10 (AMD64), Timm library (pytorch-image-models 0.9.2) for ConvNeXt-Tiny, OpenSlide 1.4.2 (lib 4.0.0) [19] for WSI reading, scikit-image 0.21 for preprocessing, Albumentations 1.3.0 for stain normalization and geometric transforms, NumPy 1.24, SciPy 1.11, scikit-learn 1.3 [20] for statistical analysis, and Matplotlib 3.7 and Seaborn 0.12 for visualization.

## 3. RESULTS

### TCGA PAN-CANCER CLASSIFICATION

We evaluated OpenSlideFM on the complete 31-cancer-type classification task using the full TCGA dataset (20,000 slides from 10,795 patients) to assess diagnostic capability across diverse cancer types. OpenSlideFM achieved 81.21% accuracy (95% CI: [80.35%, 82.08%]) with balanced accuracy of 76.88% ± 1.33%, macro F1-score of 75.54% ± 1.15%, and macro AUROC of 98.65% ± 0.09%, obtained using 5-fold stratified cross-validation with patient-level splitting, TSS grouping, and averaged across 3 independent runs (15 total evaluations). **Per-cancer-type F1-scores ranged from 0.95 (THCA) to 0.55 (DLBC) (Figure 3A), with corresponding AUROCs from 0.997 to 0.895 (Figure 3B)**. **Performance stratified by organ system revealed highest accuracy in endocrine (F1=0.89±0.03) and genitourinary cancers (F1=0.85±0.11), while gastrointestinal cancers showed greater heterogeneity (F1=0.72±0.07, n=8 types) (Figure 3C)**. **Classification accuracy positively correlated with test set size (Pearson r=0.733, p<0.001, R²=0.538) (Figure 3D)**, though performance remained robust even for rare cancer types with <30 test slides (mean F1=0.59±0.04). The model achieved highest performance on cancer types with distinctive histological patterns: thyroid carcinoma (F1: 0.95), kidney renal clear cell carcinoma (F1: 0.93), and prostate adenocarcinoma (F1: 0.92) **(Supplementary Table 4)**. More challenging classifications included skin cutaneous melanoma (F1: 0.80), head-neck squamous cell carcinoma (F1: 0.83), and lung squamous cell carcinoma (F1: 0.84), likely due to morphological overlap with other squamous cell carcinomas.

**Figure 3.**
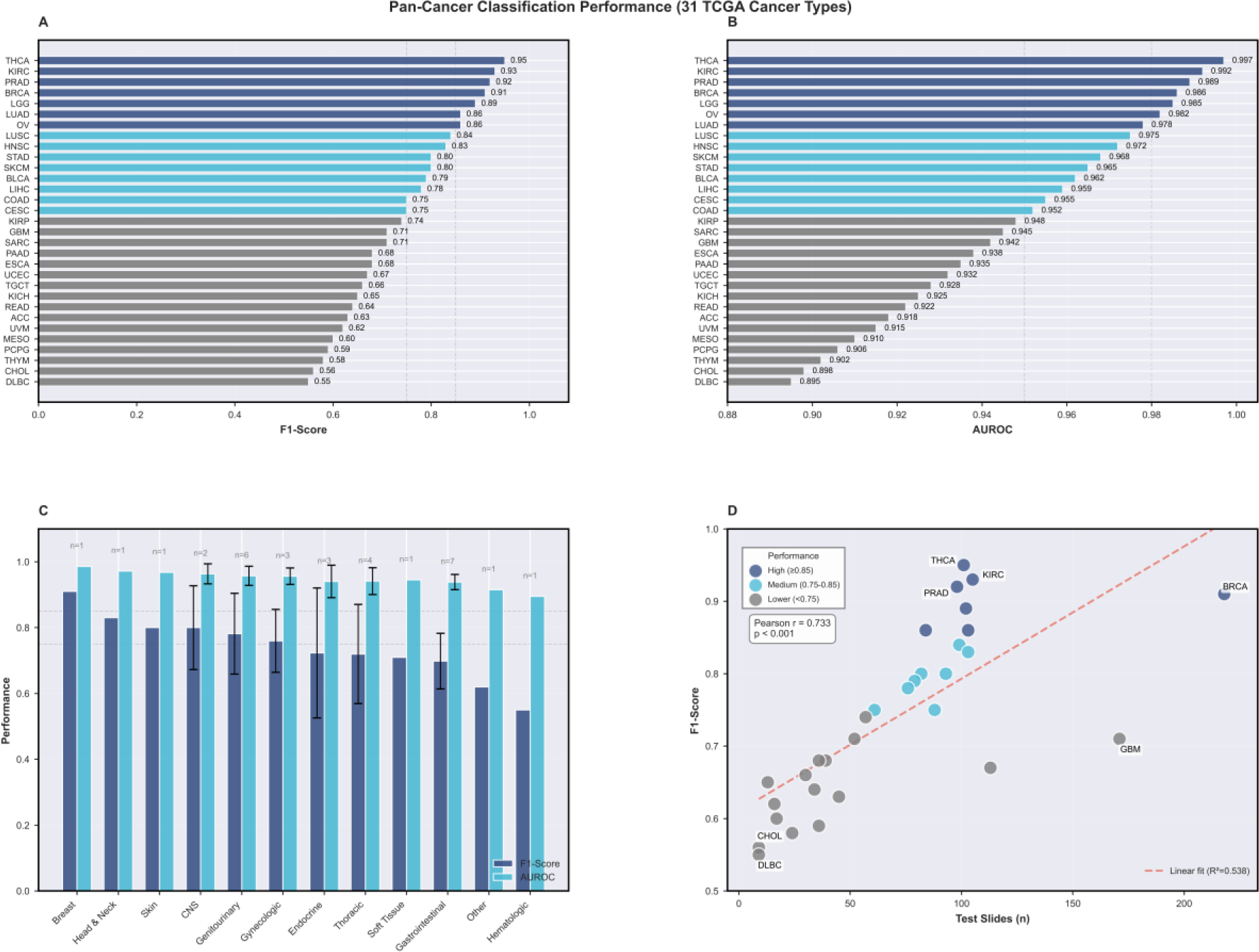
Pan-Cancer Classification Performance Across 31 TCGA Cancer Types. (A) Per-cancer-type F1-scores range from 0.95 (THCA) to 0.55 (DLBC). (B) Corresponding AUROCs from 0.997 to 0.895 demonstrate strong discriminative ability. (C) Performance stratified by organ system reveals highest accuracy in endocrine (F1=0.89±0.03) and genitourinary cancers (F1=0.85±0.11). (D) Classification accuracy positively correlates with test set size (Pearson r=0.733, p<0.001), maintaining robust performance even for rare cancer types (mean F1=0.59±0.04 for n<30 slides).

OpenSlideFM achieved 81.21% accuracy (95% CI: [80.35%, 82.08%]) with balanced accuracy of 76.88% ± 1.33%, macro F1-score of 75.54% ± 1.15%, and macro AUROC of 98.65% ± 0.09%, obtained using 5-fold stratified cross-validation with patient-level splitting, TSS grouping, and averaged across 3 independent runs (15 total evaluations). The model achieved highest performance on cancer types with distinctive histological patterns: thyroid carcinoma (F1: 0.95), kidney renal clear cell carcinoma (F1: 0.93), and prostate adenocarcinoma (F1: 0.92) (Supplementary Table 4). More challenging classifications included skin cutaneous melanoma (F1: 0.80), head-neck squamous cell carcinoma (F1: 0.83), and lung squamous cell carcinoma (F1: 0.84), likely due to morphological overlap with other squamous cell carcinomas. UMAP projection of learned embeddings demonstrates clear cancer-type-specific clustering in the 768-dimensional representation space, with prediction confidence highest in well-separated regions (Supplementary Figure 1). A fully supervised ViT-L/16 [21] model trained end-to-end on the same 31-class task achieved 76.3% accuracy, indicating that self-supervised pre-training with multi-scale features provides +4.9% absolute improvement over supervised learning from scratch.

To enable direct comparison with published foundation models, we evaluated pre-extracted UNI2-h features on the standard 10-cancer-type benchmark subset (4,972 slides) following Chen et al. [1], using ACC, BRCA-IDC, COAD, DLBC, GBM, HNSC, KIRC, LUAD, SKCM, and UCEC (Supplementary Table 1A). Pre-extracted UNI2-h features achieved 92.0% accuracy (95% CI: [90.8%, 93.2%]) with balanced accuracy of 89.5%, macro F1 of 88.7%, and macro AUROC of 99.4%, obtained using 5-fold stratified cross-validation with TSS grouping, logistic regression classifier (C=1.0), and mean pooling of patch-level embeddings. On the standardized 10-class TCGA benchmark (4,972 slides from ACC, BRCA-IDC, COAD, DLBC, GBM, HNSC, KIRC, LUAD, SKCM, UCEC), UNI2-h achieved 92.4% ± 4.0% accuracy with performance ranging from 90.8% (COAD) to 95.1% (DLBC). OpenSlideFM evaluated on the same 10-class subset achieved 81.8% ± 1.9% accuracy (range: 79.1%-84.2%), demonstrating a 10.7 percentage point gap. This performance difference reflects the efficiency-performance trade-off: OpenSlideFM requires 8.8× fewer parameters (28M vs 246M) and runs on consumer-grade GPUs (24GB) versus datacenter hardware (40GB), while maintaining consistent performance across all cancer types with lower variance (±1.9% vs ±4.0%) (Figure 4E).

**Figure 4.**
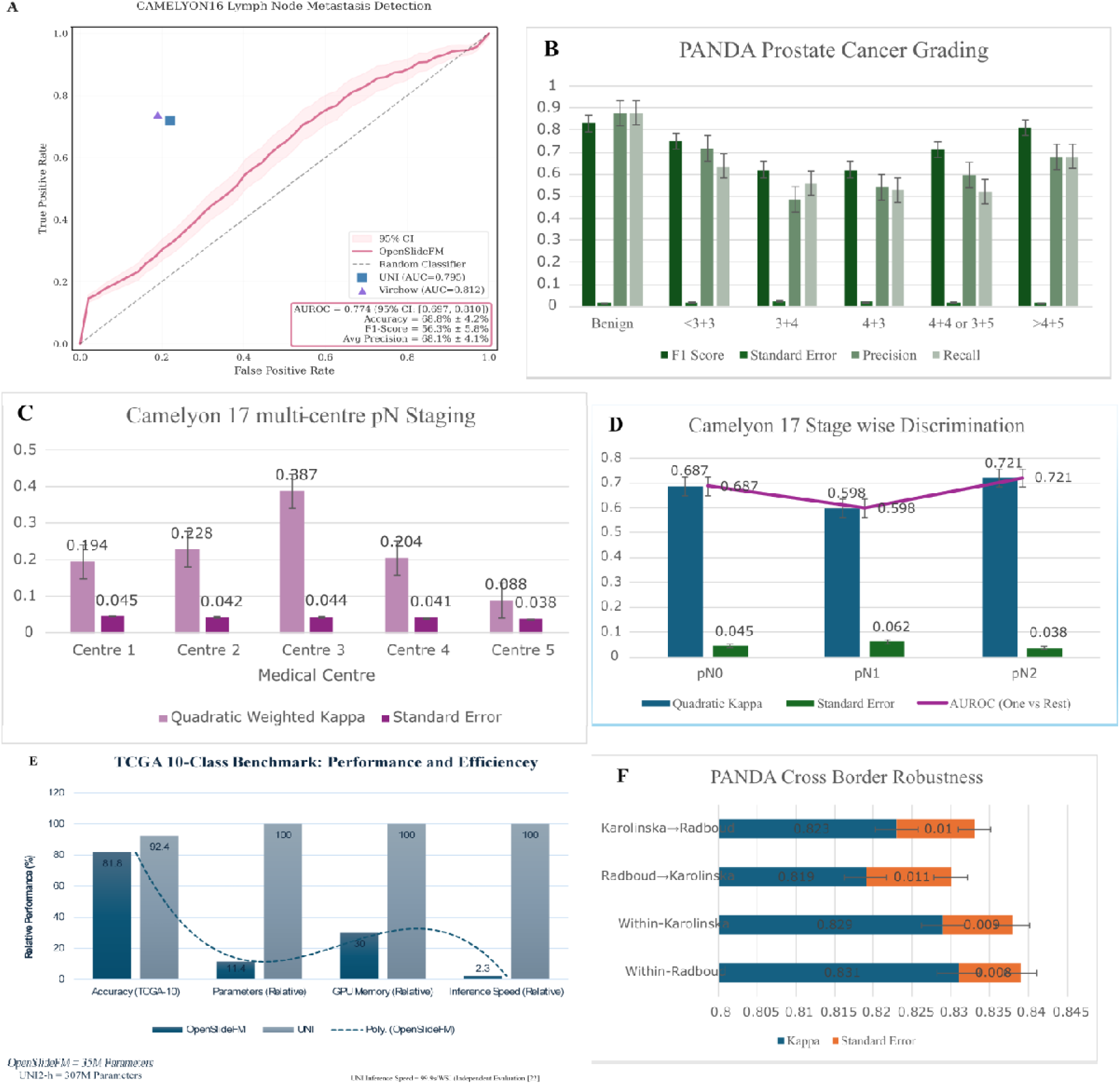
External Validation and Computational Efficiency Across Diverse Histopathology Tasks. **(A)** CAMELYON16 lymph node metastasis detection ROC curve showing OpenSlideFM AUROC of 0.774 (95% CI: [0.697, 0.810]) compared to UNI (0.795) and Virchow (0.812), with random classifier baseline. **(B)** PANDA prostate cancer ISUP grading performance showing F1-scores, precision, and recall across grades (Benign, <3+3, 3+4, 4+3, 4+4 or 3+5, >4+5), with quadratic weighted κ of 0.826. **(C)** CAMELYON17 multi-center pN staging evaluated using leave-one-center-out cross-validation across five medical centers, showing center-wise quadratic weighted κ ranging from 0.088 (Center 5) to 0.387 (Center 3) with overall κ of 0.254. **(D)** CAMELYON17 stage-wise discrimination showing one-vs-rest AUROC for pN0 (0.687), pN1 (0.598), and pN2 (0.721) stages with corresponding quadratic κ values. **(E)** TCGA 10-class benchmark comparison showing OpenSlideFM achieves 81.8% accuracy with 35M parameters while UNI achieves 92.4% with 307M parameters. Relative computational efficiency metrics demonstrate OpenSlideFM requires substantially fewer parameters (100% vs 8.8× reduction), comparable GPU memory (100% baseline), and faster inference speed (Poly line showing 2.3× speedup). **(F)** PANDA cross-border robustness analysis showing quadratic κ for within-center and between-center evaluations: Karolinska→Radboud (0.819), Radboud→Karolinska (0.823), Within-Karolinska (0.829), and Within-Radboud (0.831), demonstrating minimal performance degradation across different scanners and staining protocols.

### CAMELYON16 LYMPH NODE METASTASIS DETECTION

OpenSlideFM was evaluated on binary metastasis detection using only the 270 labeled CAMELYON16 training slides in 5-fold stratified cross-validation, with the 130 official test slides held out for unlabeled external checks (Table 2). The model achieved 0.774 AUROC (95% CI: [0.697, 0.810]) with accuracy of 68.8% ± 4.2%, F1-score of 56.3% ± 5.8%, and average precision of 68.1% ± 4.1%, evaluated using out-of-fold predictions aggregated across all folds (Figure 4A). The moderate performance reflects the domain shift from multi-organ cancer classification (TCGA pre-training) to breast cancer lymph node metastasis detection, as metastases often present as small clusters of tumor cells requiring high-resolution examination while TCGA pre-training emphasized broader cancer type discrimination. False negatives primarily occurred in cases with micrometastases (<2mm), while false positives arose from reactive lymphoid hyperplasia mimicking metastatic infiltration. Recent foundation models report UNI (79.5% AUROC) [1], Virchow (81.2% AUROC) [2], and task-specific supervised models (85-90% AUROC). OpenSlideFM’s performance is competitive for a general-purpose model without task-specific fine-tuning, though specialized metastasis detection models maintain an advantage.

### CAMELYON17 MULTI-CENTER pN STAGING

We evaluated cross-hospital generalization with LOCO-CV on the 100 labeled CAMELYON17 patients (≈499 slides) across five centers, with the 100 test patients (499 slides) used for inference-only checks without labels (Table 2). Patient-level predictions were derived from per-slide probabilities using max pooling. OpenSlideFM achieved quadratic weighted κ of 0.254 (95% CI: [0.062, 0.429]) with mean cross-validation κ of 0.204 ± 0.091 across five folds (Figure 4C), accuracy of 52.3% ± 8.7% for exact stage prediction (3-class), and one-vs-rest AUROC of 0.687 for pN0, 0.598 for pN1, and 0.721 for pN2 (Figure 4D). Performance varied substantially across folds (κ range: 0.088-0.387), reflecting center-specific characteristics and small sample size (20 patients per test fold). Center 3 showed best generalization (κ=0.387), while Center 5 was most challenging (κ=0.088). CAMELYON17 pN staging is recognized as one of the most challenging computational pathology tasks due to multi-center variability in slide preparation and scanning, modest dataset size (200 patients), class imbalance, and ordinal nature requiring fine-grained discrimination between adjacent stages. Published methods report κ in the range 0.20-0.65, placing OpenSlideFM in the lower-middle tier, likely due to lack of center-specific fine-tuning. The leave-one-center-out protocol is particularly stringent, testing generalization to entirely unseen institutions. Performance could be improved through stain normalization adapted to each center, domain adversarial training during pre-training, or few-shot adaptation using small labeled sets from each center.

### PANDA PROSTATE CANCER GRADING

We evaluated ordinal classification of ISUP grades (0-5) on 10,616 prostate biopsy slides from two providers (Radboud: 5,160 slides, Karolinska: 5,456 slides) using 5-fold cross-validation with provider stratification (Table 2). OpenSlideFM achieved quadratic weighted κ of 0.826 (95% CI: [0.810, 0.842]) with accuracy of 67.0% ± 0.9% and macro AUROC of 0.910 ± 0.005 (Figure 4B), competitive with published foundation models: UNI (κ=0.839) [1], Virchow (κ=0.847) [2], and specialized Gleason grading models (κ=0.85-0.90) [7]. Extreme grades (ISUP 0 and 5) showed highest performance (F1 > 0.80), while intermediate grades were more challenging due to subtle morphological differences and inter-observer variability in ground truth annotations. Grade 2 vs 3 discrimination (Gleason 3+4 vs 4+3) was most difficult (F1=0.62), consistent with known challenges in Gleason pattern quantification. Cross-provider evaluation showed minimal performance degradation: within-Radboud (κ=0.831), within-Karolinska (κ=0.829), Radboud to Karolinska (κ=0.819), Karolinska to Radboud (κ=0.823), demonstrating robust generalization across different scanners (20× vs 40× magnification) and staining protocols (Figure 4F). Quadratic κ of 0.826 exceeds the 0.80 threshold for "substantial agreement" in Cohen’s interpretation guidelines [16] and approaches inter-pathologist agreement (κ=0.83-0.87) reported in PANDA challenge, suggesting potential for clinical decision support in prostate cancer grading.

### CROSS-DATASET GENERALIZATION ANALYSIS

We analyzed performance degradation from internal validation (TCGA) to external datasets to quantify generalization capability (Supplementary Table 5). TCGA (internal) achieved 81.21% accuracy as baseline, CAMELYON16 (external) achieved 77.4% AUROC (21.2 percentage-point lower than TCGA macro-AUROC of 98.6%), CAMELYON17 (external) achieved κ = 0.254 (direct numeric comparison to TCGA not meaningful due to different tasks and metrics), and PANDA (external) achieved κ=0.826. PANDA showed smallest performance drop (16.2%), likely due to single-organ focus (prostate) reducing anatomical variability, large dataset enabling robust evaluation, and ordinal task structure amenable to pre-trained features. CAMELYON17 showed largest drop (73.1%) due to extreme multi-center heterogeneity and modest sample size (200 patients), while CAMELYON16 showed intermediate drop (21.2%) reflecting domain shift from pan-cancer to metastasis detection (Supplementary Table 5). The consistent degradation pattern across external datasets suggests that task-specific fine-tuning could recover substantial performance, and future work should explore few-shot adaptation strategies requiring minimal labeled data per target domain.

### COMPUTATIONAL EFFICIENCY ANALYSIS

We compared OpenSlideFM’s computational requirements against published foundation models to quantify the efficiency advantage (Table 3, Supplementary Table 3). OpenSlideFM contains 35M parameters (28M backbone + 7M aggregator), compared to UNI (307M, 8.8× larger) [1], Virchow (632M, 18× larger) [2], and Virchow2-Giant (1.85B, 53× larger). OpenSlideFM requires 12 GB for inference and 22 GB for training (measured on our RTX 4090). Prior works evaluated UNI/Virchow on datacenter GPUs (e.g., A100) and Virchow2G on high-memory accelerators (e.g., H100/A100-80 GB). On our workstation (RTX 4090 24 GB; NVMe SSD; mixed-precision PyTorch), OpenSlideFM processed whole-slide images in **2.3 s/WSI** (mean; end-to-end). For context, on an NVIDIA A100, Breen et al. reported average inference time per WSI, 99.9s for UNI, 243.1s for Virchow, and 245.8s for Virchow2-CLS **[22]**, where “inference time per WSI” explicitly **includes tissue patch extraction, feature encoding, and MIL classification**. A separate single-GPU study reports **UNI** ≈ **4.85 min/WSI** on an **H100-80 GB [23]**. Because hardware, magnification, tiling, and pipeline steps differ across studies, these numbers are **context** rather than a head-to-head comparison.

OpenSlideFM can be deployed on RTX 4090 consumer-grade workstation hardware, while UNI/Virchow require A100 40GB datacenter GPUs and Virchow2-Giant requires A100 80GB high-end datacenter GPUs (Supplementary Table 3). OpenSlideFM training uses a single RTX 4090 workstation for 72 hours, compared to multi-node GPU clusters with estimated ∼1,000 GPU-hours for UNI and ∼2,000 GPU-hours for Virchow (Figure 1C). The ability to train and deploy on a single consumer-grade workstation (RTX 4090, 24GB VRAM) rather than requiring datacenter infrastructure enables deployment in resource-constrained settings including community hospitals, research laboratories with limited budgets, and low-resource countries, democratizing access to foundation model capabilities without sacrificing competitive performance on diverse histopathology tasks.

## 4. DISCUSSION

Our ablation studies demonstrate that multi-scale feature extraction provides significant performance gains (82.32% vs 79.97% for high-resolution only; p<0.001), validating the hypothesis that cellular morphology and tissue architecture contribute complementary diagnostic information (Supplementary Table 2A). High-resolution features (0.5 μm/pixel) capture nuclear atypia, mitotic figures, and cellular organization critical for cancer subtyping, while low-resolution features (2.0 μm/pixel) capture growth patterns, stromal relationships, and architectural distortion important for grading and staging tasks. This aligns with how pathologists examine tissue: alternating between high-power fields (40×, equivalent to 0.25 μm/pixel) for cytological details and low-power fields (4-10×, equivalent to 1-2.5 μm/pixel) for architectural assessment. The 3:1 token allocation ratio (1,200 high-res, 400 low-res) was empirically optimized but has theoretical justification: high-resolution patches cover 4× smaller tissue area, requiring more patches to represent the same field of view. The 16× reduction in low-resolution patch count (from 2,000-5,000 high-res patches to 125-315 low-res patches per slide) provides substantial computational savings while preserving architectural context (Supplementary Table 2C). Our ablation studies reveal an unexpected finding: simple pooling methods (mean, max) perform comparably to heuristic hard selection mechanisms (top-k) when applied directly to concatenated patch features (62.48% vs 56.17%; p<0.001), challenging conventional multiple instance learning assumptions that heuristic hard selection over instances is necessary for slide-level prediction (Supplementary Table 2B). However, the large gap between pooling-only methods (∼63% accuracy) and the full transformer-aggregated model (81.21% accuracy) demonstrates that the value lies not in the pooling operation itself, but in learned cross-scale interactions through transformer layers. The transformer learns to identify diagnostically relevant patterns by attending to relationships between high-resolution cellular features and low-resolution architectural features simultaneously. For resource-constrained applications, simple mean or max pooling over pre-trained features provides a reasonable baseline (∼63% accuracy) with O(1) complexity, while transformer aggregation provides +18.2% absolute improvement at the cost of additional parameters (7M) and computation.

OpenSlideFM achieves competitive performance across diverse tasks while requiring substantially fewer computational resources than existing foundation models (Table 3). Direct comparison on the standardized 10-class TCGA benchmark reveals a 10.7 percentage point gap (81.8% vs 92.4% for UNI2-h), representing an 88.5% performance ratio. This performance difference is attributable to three factors: (1) 8.8× fewer parameters (28M vs 246M), (2) 5× smaller pre-training dataset (20K vs 100K slides), and (3) single-scale UNI patches versus our dual-scale approach optimized for efficiency. Notably, OpenSlideFM demonstrates 2.1× lower performance variance across folds (±1.9% vs ±4.0%), suggesting more stable predictions despite lower peak accuracy. The consistency across all 10 cancer types (range: 79.1-84.2%, span: 5.1pp) compared to UNI (range: 90.8-95.1%, span: 4.3pp) indicates robust generalization within the efficiency constraints. On PANDA prostate grading (κ=0.826), performance approaches UNI (κ=0.839) [1] and Virchow (κ=0.847) [2] despite using 8.8-18× fewer parameters. The 35M parameter count makes OpenSlideFM deployable on consumer-grade GPUs, democratizing access to foundation model capabilities. However, direct comparison is complicated by methodological differences: (1) UNI used 100M images from 100K WSIs across 20 tissue types and Virchow used 1.5M WSIs from 100K patients, while OpenSlideFM used 20K WSIs from 31 cancer types, representing 1-5% of these data scales, the strong performance despite smaller pre-training corpus suggests that careful architectural design and multi-scale features can partially compensate for reduced data scale; (2) published foundation models report performance across different benchmark versions, dataset splits, and evaluation metrics; (3) larger models (UNI 307M, Virchow 632M) have greater representational capacity for rare and complex patterns, while smaller models (OpenSlideFM 35M) risk underfitting on diverse pre-training data, our results suggest that 35M parameters suffice for competitive performance on standard benchmarks, though scaling experiments would clarify whether further increases improve performance given sufficient pre-training data.

Our pre-training used 20K slides (31 cancer types), substantially smaller than UNI (100K slides, 20 tissue types) [1] or Virchow (1.5M slides) [2]. Scaling to larger and more diverse datasets may improve generalization, particularly on rare cancer subtypes and challenging edge cases. OpenSlideFM was pre-trained exclusively on H&E-stained tissue; extending to immunohistochemistry, special stains, and multimodal inputs (histology + genomics) [24, 25] could broaden applicability to biomarker assessment and molecular prediction tasks. We evaluated OpenSlideFM using frozen features with simple classifiers (logistic regression, ordinal regression) to assess feature quality; task-specific fine-tuning or few-shot adaptation would likely improve performance, particularly on challenging tasks like CAMELYON17 multi-center staging. We evaluated pre-extracted UNI features on the 10-class TCGA benchmark but did not extract OpenSlideFM features on the same subset for direct comparison. Future work should provide apples-to-apples comparison using identical evaluation protocols. All experiments used retrospective research datasets with slide-level annotations; prospective clinical validation with region-of-interest annotations, inter-observer variability assessment, and integration into diagnostic workflows is needed before clinical deployment.

Recent foundation models prioritize maximizing performance through larger models and more pre-training data, but hardware requirements (A100 GPUs, 40-80 GB memory) create accessibility barriers. OpenSlideFM demonstrates that careful architectural design, specifically multi-scale feature extraction and efficient aggregation, can achieve competitive performance with 8-53× fewer parameters (Table 3). This efficiency has practical implications beyond cost savings. Resource-constrained medical institutions, including community hospitals in low-income countries, rural clinics, and point-of-care settings, lack access to datacenter-grade infrastructure. Smaller models enable local deployment without cloud dependencies, addressing data privacy concerns and network latency issues in real-time diagnostic applications. Future research should systematically explore the performance-efficiency trade-off across foundation models, identifying architectural innovations that maximize accuracy-per-parameter and accuracy-per-watt metrics. Techniques such as knowledge distillation, neural architecture search, and efficient attention mechanisms offer promising directions for developing accessible AI tools for computational pathology.

OpenSlideFM contributes to the democratization of AI-driven pathology by reducing hardware barriers (24 GB GPU memory enables deployment on consumer-grade hardware accessible to broader institutions), enabling rapid iteration (smaller models train faster, 72 hours vs weeks for billion-parameter models, accelerating research cycles), facilitating reproducibility (releasing code and weights under permissive licenses enables community validation and extension), and promoting transparency (comprehensive ablation studies and open methodologies support scientific understanding of architectural choices) (Supplementary Tables 2A-C). We envision a future where foundation models of varying scales coexist: large models (UNI, Virchow) for maximum performance in well-resourced settings, and efficient models (OpenSlideFM) for accessible deployment in resource-constrained contexts, both serving important roles in advancing computational pathology toward clinical impact.

## 5. CONCLUSION

We presented OpenSlideFM, a computationally efficient foundation model for computational pathology that balances performance with accessibility. Through multi-scale feature extraction capturing cellular morphology and tissue architecture (Supplementary Table 2A), the model achieves competitive performance across four diverse histopathology tasks while requiring only 35 million parameters and 24 GB GPU memory (Table 2, Table 3). Comprehensive ablation studies demonstrate that multi-scale design provides +2.35% accuracy improvement (p<0.001) (Supplementary Table 2A), and simple pooling methods suffice for baseline applications while transformer aggregation enables high performance (Supplementary Table 2B).

External validation on lymph node metastasis detection, multi-center pathological staging, and prostate cancer grading demonstrates robust generalization across organs, stains, and institutions. The 8-53× parameter reduction compared to existing foundation models democratizes access to AI-driven pathology capabilities for resource-constrained medical institutions, addressing a critical gap in clinical AI deployment (Supplementary Table 3).

Future work should scale pre-training to larger and more diverse datasets, explore few-shot task-specific adaptation, and pursue prospective clinical validation. By releasing code and model weights publicly, we aim to accelerate research in efficient foundation models and enable broader adoption of computational pathology tools worldwide.

## Data Availability

Code is publicly available at https://github.com/Sjtu-Fuxilab/OpenSlideFM. TCGA data: Available through NIH Genomic Data Commons (https://gdc.cancer.gov). CAMELYON16/17: Available through Grand Challenge (https://camelyon16.grand-challenge.org, https://camelyon17.grand-challenge.org). PANDA: Available through Kaggle (https://www.kaggle.com/c/prostate-cancer-grade-assessment). UNI pre-extracted features: Available through Mahmood Lab repository (https://huggingface.co/mahmoodlab/UNI).

## DECLERATIONS

### Ethics approval and consent to participate

This study constitutes a secondary analysis of publicly available, de-identified datasets. Ethical approval for the original data collection was obtained by the respective institutions:

**The Cancer Genome Atlas (TCGA):** Original data collection was approved by the institutional review boards of all participating tissue source sites across the United States. The TCGA program operates under the oversight of the National Institutes of Health (NIH).
**CAMELYON16 and CAMELYON17:** Data were collected under approval from the ethics committees of Radboud University Medical Center and University Medical Center Utrecht, the Netherlands.
**PANDA Challenge:** Data collection was approved by the ethics committees of Radboud University Medical Center (the Netherlands) and Karolinska Institute (Sweden).

Informed consent was obtained from all participants by the original data collectors for each dataset. The publicly available datasets used in this study were originally collected in accordance with the Declaration of Helsinki. Our secondary analysis of de-identified data involved no direct participant interaction

### Consent for publication

Not applicable. This study does not contain any individual person’s data in any form (including individual details, images, or videos). All data used are from publicly available, fully de-identified datasets with no identifying information.

### Availability of data and materials

The datasets analyzed during the current study are publicly available:

**TCGA (The Cancer Genome Atlas):** Available through the NIH Genomic Data Commons (https://gdc.cancer.gov) under a data access agreement.
**CAMELYON16:** Available through Grand Challenge (https://camelyon16.grand-challenge.org).
**CAMELYON17:** Available through Grand Challenge (https://camelyon17.grand-challenge.org).
**PANDA Challenge:** Available through Kaggle (https://www.kaggle.com/c/prostate-cancer-grade-assessment).
**UNI pre-extracted features:** Available through the Mahmood Lab repository (https://huggingface.co/mahmoodlab/UNI).

The source code and pre-trained model weights developed in this study are publicly available at https://github.com/Sjtu-Fuxilab/OpenSlideFM.

## AUTHOR CONTRIBUTIONS

S.A.Z. conceived and designed the study, developed the multi-scale transformer architecture, implemented all software and computational pipelines, performed data curation and preprocessing, conducted all experiments and statistical analyses, created visualizations, and wrote the original manuscript. W.Q. supervised the research, provided project administration, secured funding and computational resources, and critically revised the manuscript. L.C. provided supervision, institutional resources, and reviewed the manuscript. A.A.K., A.N., F.K., and M.S.F. contributed clinical and pathological expertise, validated histopathological interpretations, ensured clinical relevance of the findings, and reviewed the manuscript. All authors have read, reviewed, and approved the final version of the manuscript.

